# Trends in Violence Against Women in Morocco: An Analysis of National Surveys from 2009 to 2020

**DOI:** 10.1101/2024.07.05.24310020

**Authors:** Fatima-Ezzahraa Wafqui, Abdelhakim Yahyane, Oumnia Bouaddi, Zineb Rhajbal, Mohamed Khalis

**Affiliations:** Mohammed VI International School of Public Health, Mohammed VI University of Sciences and Health, Casablanca, Morocco; Department of Public Health, Mohammed VI Center for Research and Innovation, Rabat, Morocco; Faculty of Medicine and Pharmacy, Hassan II University, Casablanca, Morocco; Direction of Population, Ministry of Health and Social Protection, Rabat, Morocco; Faculty of Legal, Economics and Social Sciences (Souissi), Mohammed V University, Rabat, Morocco; Higher Institute of Nursing Professions and Health Techniques, Rabat, Ministry of Health and Social Protection, Rabat, Morocco

**Keywords:** Violence Against Women, Morocco, Gender-based Violence, Intimate Partner Violence, Violence

## Abstract

Violence Against Women (VAW) remains a significant challenge in Morocco despite major advances in legislative framework and policy initiatives.

**Objective:** This study aims to investigate the trends of VAW in Morocco including its forms, contexts in which it occurs and the socio-demographic characteristics of victims over a 10-year period from 2009 to 2020.

**Methods:** We conducted an online search for official publicly available, open access national reports and surveys about VAW in Morocco, published in the period between 2009-2020. We identified Five official reports. We collected data on prevalence of VAW, profiles of the victims, its forms and contexts.

**Results:** From 2009 to 2020, the overall prevalence of VAW has consistently exceeded the 50% mark with 62.80% in 2009 vs 58.2% in 2020. Although it has seen a decrease, the highest prevalence of VAW was reported among married females belonging to younger age groups, with high levels of education 75.3% in 2009 vs 62.7% in 2019. Intimate partner violence, both marital and extra-marital was the most common context where VAW occurred across the ten-year period (55% and 47.4% in 2009 respectively vs 52,5% for marital violence in 2020 and 27.7% for extra marital violence in 2019). Marital or spousal violence ranged from 38.4% among non-single women to more than 80% among victims admitted to integrated care units in 2011, 2012, and 2013. Psychological violence stood out as the most common form of violence across the years, while violence in public spaces has seen a steady decrease over the years. Electronic or cyber violence was only studied in one survey and was estimated at 4.5%.

**Conclusion:** Despite legislative advancements, VAW remains a complex issue in Morocco, with significant prevalence requiring sustained efforts to address this issue comprehensively. Education and empowerment initiatives, alongside strengthening the implementation of the law and addressing barriers to reporting, are crucial for combating VAW effectively in Morocco.

## 1. Introduction

Violence against women (VAW) is a pervasive global phenomenon with 30% of women globally having experienced some form of violence during their lifetime according to the World Health Organization WHO (1). Due to its negative consequences on the health and well-being of the victims, the 2030 UN Agenda for Sustainable Development Goals (SDGs), through their target 5.2 called for the elimination of VAW (2). Morocco, as many other countries in the world still grapples with this pervasive issue (3). Recent studies and national reports have indicated high rates of violence experienced by women across different socio-economic backgrounds and age groups. According to a report by the High Commission on Planning (HCP) in 2019, around 7.6 million out of the 13.4 million women aged 15 to 74 in Morocco have experienced at least one act of violence in the year prior to the survey (4). Various forms of violence have been observed including domestic violence, sexual assault, and intimate partner violence (4). These acts of violence lead to severe physical, psychological, and reproductive health consequences, often perpetuating cycles of trauma and inequality (5). Morocco has taken significant steps towards addressing VAW through legal reforms and policy initiatives. The enactment of Law 103-13, the “Law on the Elimination of VAW,” demonstrates the government’s commitment to combating gender-based violence. This law criminalizes various forms of violence and provides mechanisms for victims to seek legal protection and support services. Additionally, the National Strategy for the Prevention of VAW (2018-2023) aims to strengthen prevention efforts, improve healthcare responses, and enhance coordination between different sectors involved in addressing VAW (6,7). One of the most pioneering initiatives addressing VAW in Morocco was the TAMKINE program. The aim of this program was to converge the actions of all ministerial departments in the fight against VAW and allow the establishment of integrated care units for women and children victims of violence in hospitals. These units have been created at the police level and in courts with a chain of care provision that operates at the provincial and prefecture level (8–10). The initiative was then institutionalized and generalized by circular n° 1040 of June 17, 2008. The TAMKINE Program has given way to the Government Plan for Equality on the Horizon of Pairs (ICRAM, 2012-2016) whose purpose was to promote equality between women and men through eight areas of intervention, one of which concerns the promotion of equitable and equal access of women to health services (9–11).

Despite these efforts, the desired impact has not been reached (12) due to several factors related to deep-rooted patriarchal norms, traditional gender roles, and harmful cultural practices which perpetuate and normalize VAW (13). Similarly, structural factors such as poverty, limited educational opportunities, and unequal access to resources further exacerbate women’s vulnerability to violence (13). Given the significant changes over the last decade in women’s economic situation and empowerment, as well as changes of laws fostering gender equality and the protection of the rights of women in Morocco, it is important to understand the changes to the landscape of VAW in the country, across different forms and contexts, as well as the profile of survivors. Therefore, this study aims to investigate the trends of VAW in Morocco including its forms, contexts in which it occurs and the socio-demographic characteristics of victims over 10-year period from 2009 to 2020.

## 2. Materials and methods

### 2.1. Study design and data sources

We compiled data on key VAW indicators in Morocco between 2009 and 2020. Data was obtained from country-specific national surveys by the Ministry of Health and Social Protection (5,7) as well as reports by public institutions such as the High Commission for Planning (4,14), and reports by multilateral organizations operating in the field of women’s rights such as UN Women and UNFPA Morocco (15). These reports are open access and were found online through a search term strategy in French: Violence against women “Violence contre les femmes”, Ministry of Health “ Ministère de la Sante “, Morocco “Maroc”, Official Reports “Rapports officiels”. Data extraction was done through on an EXCEL sheet. Socio-demographic characteristics of participants such as age, sex and marital status were also collected and recalculated to fit the desired age group. A detailed description of each survey has been added to Supplementary File 1.

### 2.2. Definitions

We defined violence in accordance with the Declaration on the Elimination of VAW adopted by the United Nations General Assembly (1993); ““any act of gender-based violence that results in, or is likely to result in, physical, sexual or psychological harm including threats, coercion arbitrary deprivation of liberty, whether occurring in the public or private sphere” (16). The prevalence of violence was defined as the number of women reporting at least one incidence of violence during the considered reference period (a year, adult life, or lifetime). The prevalence rate was defined as the proportion of women who have experienced violence in the overall population under consideration. This population varies depending on the type of violence and the living context (or setting) where the violence occurred. Regarding the various forms and contexts of VAW, definitions are shown in Box 1 below.

#### Box 1.

**Definitions of forms and contexts of VAW**

**Physical violence:** refers to any act of physical violence against a girl or woman because of her gender. Physical violence includes but is not limited to: slapping; throwing objects that could hurt; pushing, shoving or pulling hair; hitting; kicking, biting, dragging; kicking; choking; burning; threatening with a knife, firearm or other weapons among others. (4).

**Sexual violence:** refers to any harmful or unwanted sexual behaviour imposed on a person. Sexual violence includes acts of abusive sexual contact, forced engagement in sexual acts, attempted or performed sexual acts with a woman without her consent, which may include; rape and attempted rape, sexual harassment, unwanted touching, incest etc. (4).

**Psychological violence** includes both emotional abuse and behavioral control. Emotional abuse was defined as insulting; belittling or humiliating in front of others; frightening or intimidating; and threatening to hurt. Behavioral control was defined as isolating, preventing from visiting family or friends; monitoring social interactions; ignoring and indifference; making unjustified accusations of infidelity; controlling access to health care or to education etc. (4).

**Economic violence**: occurs when a person denies his or her partner access to financial resources, usually to abuse or control her, or to isolate her or impose other negative consequences on her well-being, usually to abuse or control her, or to isolate her or impose other negative consequences on her well-being (4).

**Workplace violence:** manifested mainly in discrimination in terms of pay, promotion or training and internships compared to male colleagues, preferential treatment of male workers compared to female workers etc (4).

**Judicial violence:** refers to any unfair application of the law that favours men more at the expense of women. In the context of a marital relationship, the laws in question pertain to issues of child custody and alimony in case of divorce or separation, as well as inheritance matters following the death of a partner (4).

**Intimate Partner Violence (IPV):** includes any act of violence, whether psychological, physical, sexual, or economic, or related to the enforcement of the law, perpetrated by a husband/ex-husband, a fiancé/ex-fiancé, or an intimate partner/ex-intimate partner (4).

**Domestic violence:** refers to the exercise, or threat of exercise, by a structurally advantaged or stronger person, of physical, psychological, sexual or economic violence within a family relationship other than conjugal. This includes acts of violence committed by family relatives such as brothers, male cousins, in-laws etc. (4).

**Education and institutional violence:** refers to any form of VAW perpetrated in or around education and training establishments against female pupils or students by teachers, administrative staff, fellow students, or other outsiders in the vicinity of the establishment (4).

**Public space violence:** refers to any form of VAW in the public space, which may be perpetrated by acquaintances (friends, neighbours) or strangers, as well as by law enforcement officers or service providers in government departments or elsewhere (4).

**Electronic violence or cyber-violence:** refers to any intentional aggressive act by an individual or group of individuals (known or unknown to the victim) using electronic forms of communication. Examples include posting intimate pictures, contacting the victims’ relatives through social platforms or monitoring publications, any form of verbal abuse on chat boxes etc. (4).

### 2.3. Data Analysis

We analysed and compared the overall prevalence of VAW over the period of ten years from 2009 to 2020. The prevalence of VAW was reported by age group, place of residence, matrimonial status, educational backgrounds, type of violence (physical, sexual, psychological, economic, workplace and judicial); and context of violence (intimate partner violence, domestic, violence in public spaces, education and institutional, electronic).

## 3. Results

### 3.1. Prevalence and socio-demographic characteristics

A total of 5 official reports have been published between 2009 and 2020 and were all included in the analysis (4,5,7,14,15) (See table 1). The overall prevalence of VAW ranged from 57.0% to 62.8% over this period and the highest prevalence was reported among married women reaching 63.6% in 2009. Prevalence of VAW across age groups was reported in four surveys and was found to be higher in the youngest age group < 25 years old ranging from 70.3% in 2009 to 35.8% in 2019. The oldest age group >50 had the lowest prevalence of VAW ranging from 48.4% in 2009 to 22% in 2019. Prevalence across place of residence was available in four reports and ranged from 17 to 67.5% in urban settings and from which 11.9 to 56% in rural areas. Of these reports, 3 reported slightly higher prevalence rates in urban areas and one showed an opposite trend. Regarding educational level, prevalence of VAW was higher among women with higher education levels ranging from 19.4% to 75.3% among women with university education, and from 13.4% to 56.8% among unschooled women. Concerning marital status, married women experienced more violence compared to single women in two surveys (52.1% and 44.02% vs 47% and 27% respectively) whereas single women experienced more violence in one survey (67.5% vs 63.6% married in 2009).

**Table 1:**
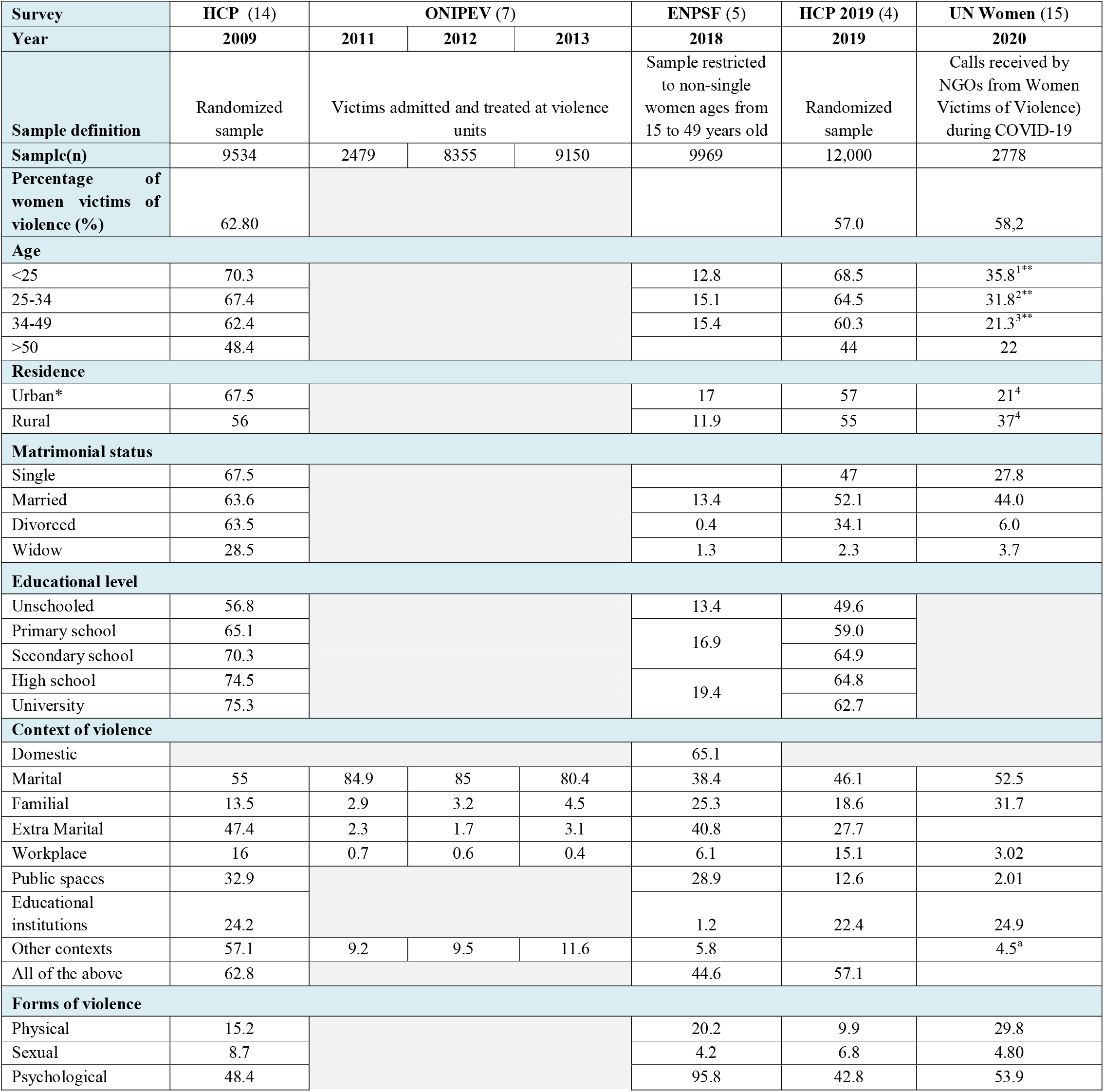
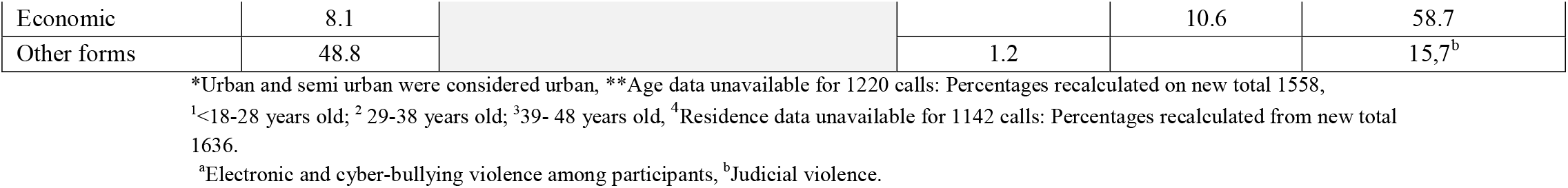
Prevalence, form and contexts of VAW in Morocco 2009 - 2020.

### 3.2. Forms of violence against women

Six forms of VAW were reported in the surveys: domestic, physical, sexual, psychological, economic and cyberviolence. Of these, psychological violence was the most prevalent over the period 2009 – 2020 ranging from 42.8% to 95.8% across the years. In 2020, economic violence was the most prevalent form (58.74%). The most uncommon form of violence was sexual violence ranging between 4.2 and 8.7% across the years. Judicial violence was reported in one survey 15.76% in 2020.

### 3.3. Contexts of violence against women

Across all surveys, marital (spousal) violence was the most prevalent context ranging from 38.4% among non-single women to more than 80% among victims admitted to integrated care units in 2011, 2012, and 2013. The prevalence of extra-marital violence was also high in 3 surveys, ranging from 27.7% to 47.4%. Regarding violence in public spaces, we noted a steady decrease. The highest prevalence was reported in 2009 (32.9%) whereas the lowest prevalence was recorded in the most recent survey (2.01% in 2020). In educational settings, the prevalence of VAW was above 22% in 2009, 2019 and 2020 whereas the lowest percentage was recorded in 2018 (1.2%). Electronic violence was reported by one survey in 2020 and showed that 4.57 % of the study population experienced at least one episode of cyberviolence in the 12 months preceding the survey.

## 4. Discussion

The aim of this study was to describe the prevalence of VAW in Morocco between 2009 and 2020 reported by national surveys and to describe the forms and contexts of the violence, and socio-demographic characteristics of the victims. Over this period, we found that the overall prevalence of VAW exceeded 50%. The highest prevalence of VAW was reported among married females belonging to younger age groups and with high levels of education. The most common contexts where VAW occurred were in conjugal or extra-marital contexts and were perpetrated by an intimate partner (spouse or otherwise). In addition, public space violence has seen a notable decrease over the years and psychological violence stood out as the most common form of violence across the years.

The findings in this study are consistent with trends in other countries in the region and in the Arab world, which highlight high level of VAW. In Turkey, a staggering 97% of women reported having experienced both physical and psychological abuse from their partners and relatives at least once in their lifetime, according to a study report published in 2022 (19). Similarly, Egypt has seen a high number of domestic violence cases in 2005, with women suffering severe consequences, including death (20–23). In Bahrain, 30% to 40% of attempted suicides are by foreign female domestic workers subjected to verbal, physical, and/or sexual abuse (24). Alternatively, studies in Pakistan, for instance show higher rates of violence reaching 70-90% of women who experienced domestic violence in 2008 (17,18).

In this study, we found a high a prevalence of IPV among women in Morocco across the years with marital violence accounting for almost half of the reported cases since 2009. In addition, married women consistently experienced higher rates of VAW compared to single or divorced women. Many studies and official reports reveal that IPV is prevalent in the MENA region (10). According to the World Bank, at least 35% of women in MENA have experienced some form of violence by an intimate partner during their lifetime, making the region the second highest in the world regarding VAW prevalence (25). In Algeria, a national report showed that one in every ten women aged 19–64 is battered almost every day (26). The presence of violence in some Arab households is often explained by deep-seated discriminatory cultural beliefs and attitudes which lead to the normalization of violence committed by the husband (27,28). This normalization may be further emphasized as women in the Arab world begin to perceive abuse as a regular aspect of their lives. Many women do not consider these acts as violence per say, which leads to them underreporting occurrences of VAW. While there is limited data on this in Morocco, a survey conducted in 2022 by “Afro Barometer” reported that in nearly half of the cases, women in rural and low socio-economic background, believe it is justified for a husband to physically harm his wife when discussing opinions (51%), going out without informing him (50%), neglecting the children (49%), refusing sexual relations (43%), or burning food (24%) (29,30). Many men in the MENA region also perceive the legitimacy of violence against intimate partners based on societal expectations of male dominance within households and relationships (31,32). To combat VAW, many studies highlight the importance of education at home and in schools and of empowering women and girls with knowledge about their rights and how to respond to violence is crucial (16,33,34). Noteworthy efforts to combat VAW in Morocco involve various governmental institutions, sectors and initiatives. Tamkin Initiative or ‘Empower’ for example, is a governmental multi-sectoral joint programme that offers legal and economic support to abused women through 52 counselling centres established in 2010 (35). A number of Moroccan NGO’s have also been vocal about this issue and are actively engaged in empowering women in Morocco, from offering legal assistance for women to defend themselves in court to advocating for the amendment of legislative laws (33,36). Up to 2019, most initiatives haven’t been able to drastically impact the prevalence of VAW in Morocco (33,35) showcasing the imminent need for intensified and versified efforts on different fronts. The new national policy to combat VAW by 2030 that was adopted in June 2021, aimed to counteract the stagnating trends (37). Since then, a number of anti-violence mechanisms have been implemented and existing programs have been strengthened. Examples of these initiatives are the adoption of a national strategy on human trafficking, the revamping of the national commission for the care of women victims of violence, as well as the national observatory of VAW, along the increases in budgets and staffing for existing counselling units for women victims of violence etc.(37)

This study, showed that low educational background appears to be a protective factor, as women with higher education levels also experience higher levels of VAW. This counterintuitive finding shows the complex relationship between education and VAW in the Moroccan context and also points to possible underreporting trends among lower educational background groups. Other studies in the literature also show similar trends whereby higher education is associated with lower reporting of occurrences of VAW (31). Nevertheless, regardless of educational background, reporting of VAW is a major issue that hinders understanding of the state of VAW in Morocco and globally (33,38). Under-reporting in Morocco may be due to societal attitudes and normalization, particularly within the context of IPV, but also due to general perceptions about the non-enforcement of the laws or lack of knowledge about the laws in general (11,39,40). Response to cases of victims who do report the occurrence of VAW also remains inadequate and may discourage victims from seeking legal help and has been subject to criticism by civil society actors. While there is limited data on this, a report by the NGO Mobilising for Rights Associated (MRA) reported that of 92,247 women who sought help at the VAW units at First Instance or Appeals courts, only 21,588 (23%) benefited from legal aid and only 4,233 (4.6%) resulted in a court hearing (34). Thus, there is a need to strengthen legal frameworks and legislative advances in combatting VAW and address barriers to reporting in Morocco. It is argued that even investments in the education of women and increased economic opportunities may not be effective in the absence of supportive state response to violence (34). Failure to enforce the laws and impunity of perpetrators is increasingly gathering media attention and undermines trust in the justice system and may contribute to perpetuating a cycle of violence and impunity. Thus, efforts to strengthen the implementation of law must be coupled with public awareness and education campaigns to change societal attitudes towards violence and promote a culture of accountability and respect for human rights, while also putting in place robust reporting mechanisms to encourage victims to come forward and to reassure the public of the commitment to justice and safety.

While this study on the prevalence of VAW across a ten-year period in Morocco is informative, some limitations to our analysis need to be considered. The study used data from national surveys with differences in the definition of VAW, types of respondents, and methods. This means that the study relies on potentially inconsistent and non-homogeneous data sources with varying methodologies, leading to difficulties in making robust comparisons. Underreporting due to cultural stigmas likely results in prevalence rates underestimating the findings which is a universal challenge when studying VAW. Finally, while the study enables us to observe the overall trend and changes in VAW, it does not allow us to identify explanatory factors for the observed changes.

## Conclusions

VAW remains a significant issue in Morocco with far-reaching consequences, despite significant legislative advances. There is a crucial need for intensified efforts to improve the implementation and enforcement of existing legal frameworks. Furthermore, there is a pressing need for future research aimed at comprehensively understanding the existing and persisting barriers to reporting violence experienced by women across diverse forms and settings, and across different demographics. This nuanced understanding will facilitate the development of evidence-based policies and initiatives to effectively address violence in various contexts, and may also be used to produce meaningful amendments to the current legal framework.

## Data Availability

All data produced in the present study are available upon reasonable request to the authors.

