## Supplemental Table 2 for "Trends in Violence Against Women in Morocco: An Analysis of National Surveys from 2009 to 2020"

**Supplementary File 1**

| **Qualitative analysis report on violence against women during lockdown in Morocco** | **UN Women** | 2020 | Over 20 civil society organizations partnered to create a qualitative analysis report on calls received by their women's support centers during the COVID-19 lockdown. The coordination was supported by ONU Femmes as part of the "Enhanced Prevention and Intervention in Cases of Violence Against Women in Morocco" project, funded by the Canadian government. Scope of the Report: The report covers information gathered from women and girls who contacted the support centers between March 20 and May 30, 2020. It addresses various forms of violence (physical, psychological, economic, sexual, legal) across different contexts (marital, familial, digital, institutional, public, professional). The goal is to provide insights into the experiences of these women, highlighting obstacles in accessing services based on the analysis of 2,778 calls out of 4,768 received by the organizations. |
| --- | --- | --- | --- |
| **National Survey on Violence against Women and Men 2019** | **HCP** | 2019 | The 2019 National Survey on the Prevalence of Violence against Women is a continuation of the previous survey conducted in 2009, in line with the recommendations of the United Nations Statistics Division (UNSD). Compared to the 2009 survey, the 2019 survey aims to update data on gender- based violence and broaden its scope of investigation and analysis. Indeed, the new survey not only targets a larger population of women and adolescent girls (aged 15 to 74, instead of 18-64 years for the 2009 survey), but it also considers other dimensions of this phenomenon, including electronic violence (also known as cyber-violence), the representations and perceptions of victims of violence from this phenomenon, as well as its its social and economic impacts on households. The approach taken in this survey is quantitative. It is based on direct interviews, through questionnaires, with a sample of 12,000 girls and women and 3,000 boys and men, aged 15 to 74, representing the various social strata and regions of the country. |
| **National population and family health survey** | **Moroccan Ministry of Health** | 2018 | The Ministry of the Family, Solidarity, Equality and Social Development launched the second national survey on the proliferation of violence against women at the end of 2017. Its objectives were not only to provide new and precise data on the proliferation of this phenomenon and to study the different reasons and factors leading to the exercising of violent acts based on the social gender in view of social and behavioural transformations experienced by Moroccan society, but also to open a new era for the promotion and protection of women's rights through a review of the national strategy for combating violence against women and make it highly targeted as well as various regional and local programs. The most important objectives of this national study are to: -Define the rate of proliferation of violence against women at the national level, and its prevalence as per the environment and the forms listed in Law 103.13 on Combating Violence against Women; -Define the characteristics of women victims of violence, their socio-economic environment, and the characteristics of the perpetrators of violence, and their socio-economic environment; -Use the results of national research to define and inspect initiatives capable of eliminating this phenomenon. The national survey conducted between January and March 2019 involved 13,543 female subjects, aged between 18 and 64, from all over the Kingdom. The model sample prepared in 2015 by the High Commissioner for Planning for the total census of the population was used, based on the use of probability methodology. |
| **(ONIPEV)**  **Information system for women and children’s victims of violence** | **Population Directorate, Moroccan Ministry of Health** | 2014 | Data from the information system of the Ministry of Health show that women victims of violence (FVV) who have used the services of the Integrated Units for the Care of Women and Children Victims of Violence (UIPFEVV) located in different hospital structures in the country (excluding university hospitals) |
| **National Survey on the prevalence of violence against women** | **HCP** | 2009 | According to the survey data, out of a population of 9.5 million women aged 18-64, nearly 6 million, or 62.8 per cent, had experienced some form of violence in the 12 months preceding the survey, 3.8 million in urban areas and 2.2 million in rural areas. |
